# Comparison of outcomes of Long versus Short Cephalomedullary nails for the fixation of intertrochanteric femur fractures: A Protocol for Systematic Review and Meta-analysis

**DOI:** 10.1101/2023.01.21.23284859

**Authors:** Rajesh Kumar Rajnish, Amit Srivastava, Prasoon Kumar, Sandeep Kumar Yadav, Siddhartha Sharma, Rehan Ul Haq, Aditya Nath Aggarwal

**Author notes:** **Correspondence to:** Dr. Rajesh Kumar Rajnish. Department of Orthopedics, All India Institute of Medical Sciences, Jodhpur, India. 342005. **Amendments:** None. **Contributions:** ANA, AS, RUH, and PK contributed in protocol conceptualization and gave critical inputs to formulate the protocol. RKR and SS planned the search strategy for literature. SKY and RKR wrote the manuscript, and RKR proofread it. Finally, this review protocol was approved by all the authors as the final version of the protocol for publication.

## Abstract

**Background:** The incidence of intertrochanteric femur fracture (IFF) in the elderly has increased with increased life expectancy globally. These fractures require surgeries at the earliest to bring them out of bed and minimize the complications of recumbency, like decubitus ulcers, decreased cardiopulmonary reserves, and thromboembolic events. Dynamic hip screws and Cephalomedullary nails (CMN) are both considered adequate for the internal fixation of the stable IFF with comparable stability and outcomes. However, CMNs are considered to have better results in unstable IFF fixation.

**Objective:** To compare the outcomes of internal fixation of short CMN versus long CMN of IFF in the elderly by analyzing the evidence from the current literature.

**Methods:** A systematic review and meta-analysis will be performed in accordance with the PRISMA guidelines. A primary search of Medline, Embase, Scopus, and Cochrane Library databases will be conducted using a pre-defined search strategy. The studies of any design in the English language will be included, which have compared the outcomes of the internal fixation of short CMN versus long CMN of the IFF and reported at least one primary or secondary outcome of interest.

Studies that did not compare the outcomes of the internal fixation of short CMN versus long CMN of the IFF, conference abstracts, posters, case reports, book chapters, technical tips, review articles, biomechanical studies, cadaveric studies, and the articles not in the English language will be excluded.

Both qualitative and quantitative analyses will be performed. A qualitative analysis will be performed using appropriate tables and diagrams. Wherever feasible, quantitative analysis will be done with the appropriate software. The risk-of-bias assessment for non-randomized comparative studies will be done using the MINORS tool, and the Cochrane Collaboration’s risk-of-bias tool will be used for randomized control trials (RCT).

## 1. Background

The incidence of IFF in the elderly has increased with increased life expectancy globally. [1] The majority of elderly patients have some or the other medical co-morbidities, which can potentially increase the associated morbidity and mortality in these patients. [2] These fractures require surgeries at the earliest to bring them out of bed and minimize the complications of recumbency, like decubitus ulcers, decreased cardiopulmonary reserves, and thromboembolic events. It has been documented that surgery delays for hip fractures lead to increased mortality in the elderly. [3]

Both dynamic hip screws (DHS) and CMN are considered adequate for the fixation of the stable IFF with comparable stability and outcomes. [4] However, the adequacy of DHS in the fixation of unstable IFF is relatively inferior to CMN [5] and often leads to complications like loss of reduction, implant failure, or lag screw cut out [6]. The CMNs have shown better results in unstable IFF fixation and are considered more reliable. [7]

In terms of design, although both short and long CMNs are suitable for the fixation of IFF, the use of short CMNs has reported advantages of shorter surgical duration, lesser intraoperative blood loss, and blood transfusion. [8] On the other hand, long CMNs have the theoretical advantage of minimizing stress concentration near the distal end of the nail, hence potentially decreasing the occurrence of secondary fractures of the shaft femur [9]. However, the superiority of one design over the other has been a matter of debate despite multiple studies in the literature. [8-10]

## 2. Need for review

There is a lack of clarity regarding the superiority of short CMN over the long CMN fixation of IFF in elderly patients. There are published reviews on the current topic, however, with a limited number of articles and outcomes. [8-11] Hence, further analysis is needed for the evidence of the superiority of one implant over another, including a maximum number of published studies. Therefore, the current review aims to perform a systematic review and meta-analysis of the current literature to compare the outcomes of IFF internal fixation with short CMN versus long CMN.

## 3. Objectives

### Primary Objectives

i. To compare the primary outcomes like ipsilateral shaft femur re-fractures, and mortality following internal fixation of IFF with short CMN versus long CMN.

### Secondary Objective

i. Additionally, to compare implant-related complications, overall complications, intra-operative blood loss, duration of surgery, re-operation rates, blood transfusion required, lengths of hospital stay, and fluoroscopy time.

## 4. PICO framework for the study

i. *Participants:* Human adults with intertrochanteric femur fractures
ii. *Intervention*: Internal fixation with short CMN
iii. *Control*: Internal fixation with long CMN
iv. *Outcomes*: the primary outcome will be ipsilateral shaft femur re-fractures and mortality. Secondary outcomes will be implant-related complications, overall complications, re-operation rates, duration of surgery, intra-operative blood loss, blood transfusion required, lengths of hospital stay, and fluoroscopy time.

## 5. Methods

This systematic review and meta-analysis will be performed in accordance with the Preferred Reporting Items for Systematic Reviews and Meta-analysis (PRISMA) guidelines. [12] A protocol for the review was formulated and registered to the PROSPERO database vide registration number CRD42023390520.

i. Review Protocol: A protocol for the review will be formulated in accordance with the PRISMA-P guidelines. (Appendix I)
ii. Eligibility Criteria: The studies of any design in the English language will be included that compare the outcomes of the internal fixation of short CMN versus long CMN of internal fixation of IFF and report at least one primary or secondary outcome of the review. Studies that do not compare the outcomes of the internal fixation of short CMN versus long CMN of the IFF, case reports, conference abstracts, posters, book chapters, review articles, biomechanical studies, technical tips, cadaveric studies, and articles not in the English language will be excluded.
iii. Information Sources & Literature search: A primary literature search will be conducted on the Medline, Embase, Scopus, and Cochrane Library databases, using pre-defined search strings of keywords *“(intertrochanteric AND fracture OR (pertrochanteric AND fracture) OR ‘hip fracture’) AND ‘*intramedullary *nail’ OR ‘cephalomedullary nail’ OR ‘cephalomedullary nail*.*”* A manual secondary search of the bibliography of the full text of all included articles and relevant review articles will be conducted. Articles published from the year 2000 till the date of the search will be included.
iv. Study Selection: All the identified articles will be screened through titles and abstracts for eligibility independently by three authors. After initial screening, full texts of all selected articles will be obtained. Eligible articles will be sorted as per the prespecified inclusion and exclusion criteria. The reasons for excluding those articles for which full text was obtained will be documented. Any discrepancies in the article selection process will be resolved by mutual agreement.
v. Data Collection & Data Items: Data will be extracted on pre-formed data collection forms by two authors independently, and a third author will cross-check the data for accuracy. Baseline data items will include:
  1. Name of authors and year of publication
  2. Number of patients/cases
  3. Study design
  4. Type of CMN used
  5. Number of patients in each group
  6. Mean age
  7. Gender ratio
  8. Fracture classification
  9. Mean follow up
  10. Primary and Secondary outcomes
vi. Outcome Measures: The following outcome measures will be evaluated; however, addition and/or modifications will be made if needed: the primary outcomes of interest will be ipsilateral shaft femur re-fractures, and mortality. The secondary outcomes of interest will be implant-related complications, overall complications, re-operation rates, duration of surgery, intra-operative blood loss, blood transfusion required, lengths of hospital stay, and fluoroscopy time.
vii. Data Analysis and Synthesis: A qualitative data synthesis will be performed with appropriate tables and data visualization diagrams. The quantitative synthesis will be performed if ≥ 2 studies included in this review reported the values of either the primary or secondary outcomes of interest. To describe the measure of treatment effects, the mean difference will be used for continuous variables, and the odds ratio will be used for dichotomous variables. All the results will be expressed along with 95% confidence intervals. Forest plots will be made to visualize the results in diagrammatic representation. The statistical heterogeneity will be determined by using the I-square test. Reasons for clinical heterogeneity, if any, will be explored. If the heterogeneity is low (I-square value near 0%) fixed-effects model otherwise, the random effects model will be used. If possible, subgroup analysis will be performed. Publication bias will be estimated and will be shown with a funnel plot using one of the primary outcomes. Meta-analysis will be performed by using Review Manager Software version 5.4. [13]
viii. Assessment of Risk of Bias: The risk-of-bias assessment will be done using the MINORS tool for the non-randomized comparative studies [14], and the Cochrane Collaboration’s risk-of-bias tool [15] will be used for randomized control trials.

## Data Availability

Yes

---

**APPENDIX 1:**
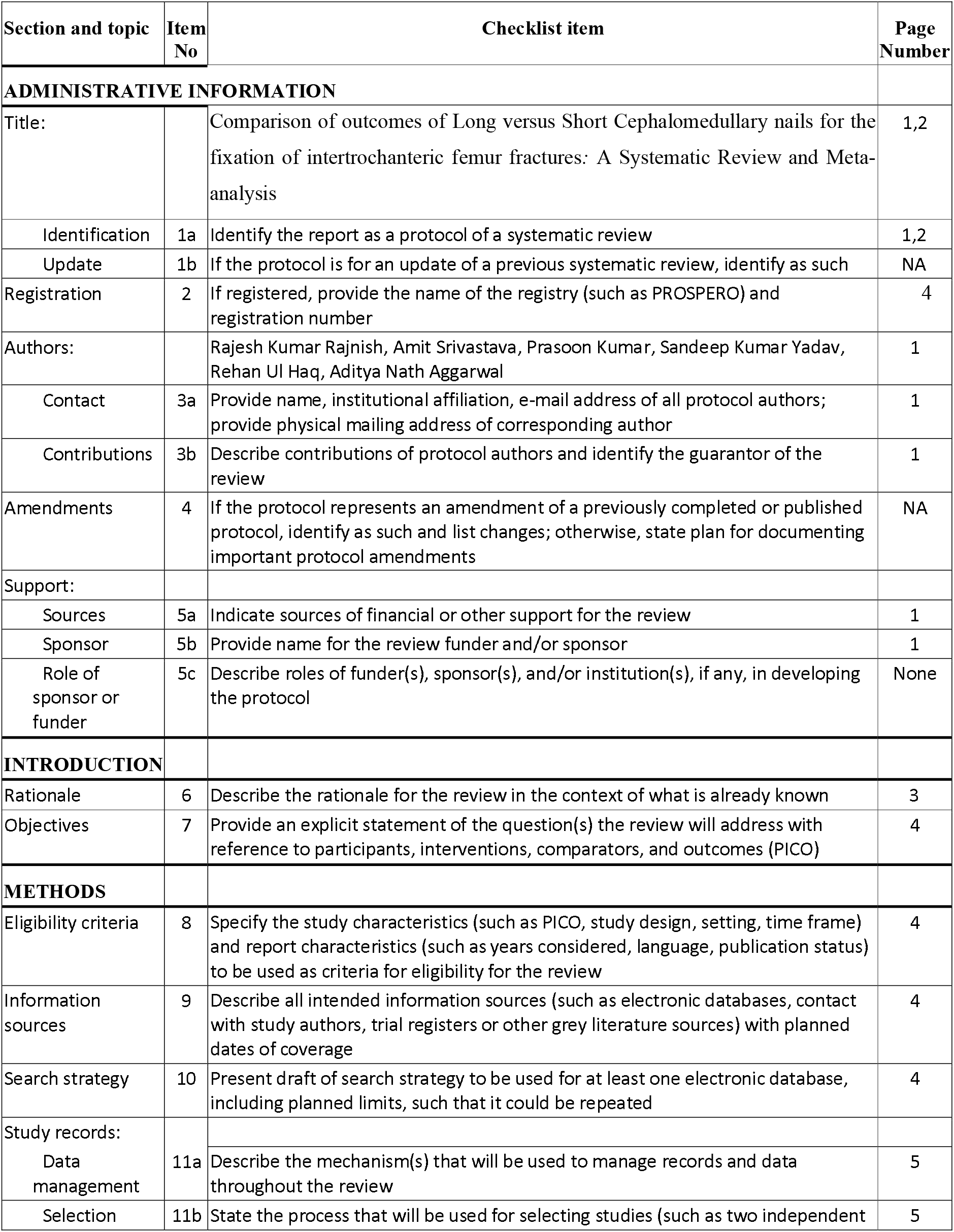
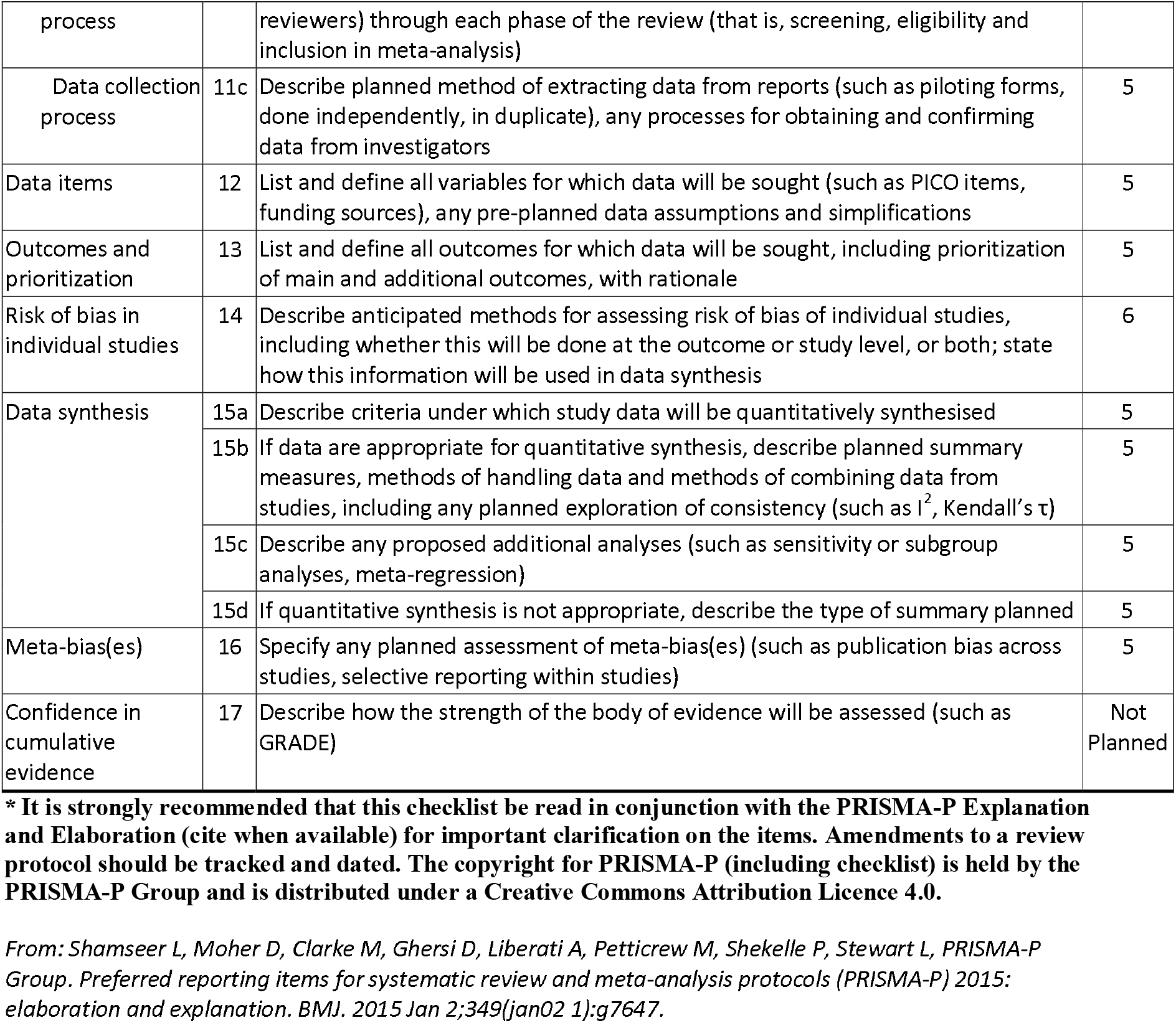
PRISMA-P (Preferred Reporting Items for Systematic review and Meta-Analysis Protocols) 2015 checklist: recommended items to address in a systematic review protocol*

